# A considerable proportion of individuals with asymptomatic SARS-CoV-2 infection in Tibetan population

**DOI:** 10.1101/2020.03.27.20043836

**Authors:** Huan Song, Jun Xiao, Jiajun Qiu, Jin Yin, Huazhen Yang, Rui Shi, Wei Zhang

## Abstract

Severe acute respiratory syndrome coronavirus 2 (SARS-Cov-2) quickly became a major epidemic threat in the whole China. We analysed SARS-Cov-2 infected cases from Tibetan Autonomous Prefecture, and noted divergent characteristics of these Tibetans infected cases compared to Han Chinese, characterizing by a considerable proportion of asymptomatic carriers (21.7%), and few symptomatic patients with initial symptom of fever (7.7%). Here, we did a descriptive study on clinical characteristics of 18 asymptomatic individuals with SARS-CoV-2 infection. The median age of these asymptomatic carriers was 31 years and one third of them were students, aged under 20 years. Notably, some of asymptomatic carriers had recognizable changes in radiological and laboratory indexes. Our finding indicates a potentially big number of SARS-CoV-2 asymptomatic carriers in prevalent area, highlighting a necessity of screening individuals with close contact of infected patients, for a better control on the spread of SARS-CoV-2 infection.

---

In December 2019, the infection of the severe acute respiratory syndrome coronavirus 2 (SARS-Cov-2) was initially emerged in Wuhan. It quickly became a major epidemic threat in the whole China[1]. Like other parts of China, Sichuan is a province affected by imported cases, and had its first confirmed 2019 coronavirus disease (COVID-19) patient reported on 21st January. Later, a county named Daofu in Ganzi Tibetan Autonomous Prefecture in western Sichuan province experienced an outbreak of SARS-Cov-2 infection in February. Particularly, Ganzi is renowned as a hypoxic plateau area and the population there is made up of mainly Tibetans. Therefore, patients in Ganzi by nature have different geographic and ethnic characteristics compared to patients in many other regions of China. Indeed, in our previous analyses of 83 cases with polymerase chain reaction (PCR)-confirmed SARS-Cov-2 infection collected from a local hospital, we noted divergent characteristics of Tibetans infected cases, characterizing by a considerable proportion of asymptomatic carriers (18/83, 21.7%), and few symptomatic patients with initial symptom of fever (5/65, 7.7%). In present research, we did a descriptive study on clinical characteristics of 18 asymptomatic individuals with SARS-CoV-2 infection.

## Methods

We conducted a retrospective, single-centre study based on the People’s Hospital of Daofu county. More than 90% of locally confirmed infected cases were recruited between 26th Jan and 28th February 2020. We defined asymptotic infected cases as the ones who got positive results on PCR test for SARS-Cov-2 infection but without any clinical symptoms during the whole surveillance period. Information on epidemiological, demographic characteristics, clinical features, treatment, and outcome were extracted from patients’ medical records. Clinical data were followed up until 6th March, 2020. The study was approved by West China Hospital Ethics Committee (reference no.2020-190) and written informed consent was obtained from patients in People’s Hospital of Daofu county before enrolment.

## Results

18 (18/83, 21.7%) individuals were identified as asymptomatic carriers, with a predominant distribution of males (61.0%) (Table 1). The median age was 31 years and one third of the asymptomatic individuals were students, aged under 20 years. Only 1 infected people (5.6%) had pre-existed chronic illness. Epidemiological data showed all the infection were caused by community transmission. Notably, some of asymptomatic carriers had recognizable changes in radiological and laboratory indexes. 7(38.9%) cases had abnormalities in chest computed tomography (CT), representing as unilateral or bilateral patchy shadowing or/and ground-glass opacity (Table 1). 9(50.0%), 7(38.8%), and 15(83.3%) cases obtained abnormal results in tests of blood routine, coagulation function, and blood biochemistry, respectively (details shown in Table 2). All asymptomatic carriers received antiviral treatment and were charged at the time of data analysis. The average duration of hospital stay was 13 days.

**Table 1.** Demographic, epidemiological, and clinic characteristics of 18 asymptomatic individuals with SARS-CoV-2 infection in Ganzi area, China.

|  | Asymptomatic infected<br>individuals (n=18), n (%) |  | Asymptomatic infected<br>individuals (n=18), n (%) |
| --- | --- | --- | --- |
| **Age, years** |  | **Type of infection** |  |
| Mean (SD) | 29.67 (13.93) | Imported cases | 0 (0) |
| Median (Rang) | 31 (6-51) | Local cases | 18 (100.00) |
| 6-20 years | 6 (33.33) | **Main findings from chest CT** |  |
| 21-51 years | 12 (66.67) | Unilateral pneumonia | 5 (27.78) |
| **Sex** |  | Bilateral pneumonia | 2 (11.11) |
| Men | 11 (61.11) | Multiple mottling | 4 (22.22) |
| Women | 7 (38.89) | Ground-glass opacity | 4 (22.22) |
| **Occupation** |  | **Treatment** |  |
| Agricultural worker | 11(61.11) | Oxygen therapy | 9 (50.00) |
| Self-employed | 0 (0) | Mechanical ventilation | 0 (0) |
| Employee | 1 (5.56) | CRRT | 0 (0) |
| Buddhist | 0 (0) | ECMO | 0 (0) |
| Students | 6 (33.33) | Antibiotic treatment | 0 (0) |
| **Chronic medical illness** |  | Antifungal treatment | 0 (0) |
| Any | 1 (5.56) | Antiviral treatment | 18 (100.00) |
| Cardiovascular diseases | 0 (0) | Glucocorticoids | 0 (0) |
| Respiratory system diseases | 0 (0) | Intravenous immunoglobulin<br>therapy | 0 (0) |
| Digestive system diseases | 1 (5.56) | **Clinical outcome** |  |
| Endocrine system diseases | 0 (0) | Discharged | 18 (100) |
| Cancers | 0 (0) | **Days from admission to**<br>**discharge, mean (SD)** | 13.39 (2.64) |
| Nervous system diseases | 0 (0) |  |  |

SD, standard deviation.

**Table 2.** Laboratory results of 18 asymptomatic individuals with SARS-CoV-2 infection in Ganzi area, China.

|  | Asymptomatic infected individuals |
| --- | --- |
| Results of laboratory tests and their changes, if any | (N=18),<br>mean (SD) or n (%)* |
| **Blood routine** |  |
| Leucocytes (x 10<sup>9</sup> per L; normal range 3.5-9.5) | 6.92(2.06) |
| Increased | 2(11.11) |
| Neutrophils (x 10<sup>9</sup>per L; normal range 1.8-6.3) | 4.82(1.76) |
| Increased | 3(16.67) |
| Lymphocytes (x 10<sup>9</sup> per L; normal range 1.1-3.2) | 1.66(0.63) |
| Decreased | 3(16.67) |
| Platelets (x 10<sup>9</sup> per L; normal range 100.0-300.0) | 158.39(52.93) |
| Decreased | 2(11.11) |
| Hemoglobin (g/L; normal range 130.0-175.0) | 152.11(19.94) |
| Increased | 2(11.11) |
| Decreased | 1(5.56) |
| **Coagulation function** |  |
| Activated partial thromboplastin time (s; normal range 22.0-38.0) | 27.23(3.40) |
| Decreased | 1 (5.56) |
| Prothrombin time (s; normal range 10.0-15.0) | 13.74(1.75) |
| Increased | 4 (22.2) |
| Fibrinogen (g/L; normal range 2.0-4.0) | 2.25(0.41) |
| Decreased | 6 (33.33) |
| **Blood biochemistry** |  |
| Albumin (g/L; normal range 35.0-55.0) | 46.18(3.10) |
| Alanine aminotransferase (U/L; normal range 0.0-40.0) | 53.78(48.44) |
| Increased | 8 (38.89) |
| Aspartate aminotransferase (U/L; normal range 0.0-40.0) | 39.44(27.83) |
| Increased | 6(33.33) |
| Total bilirubin (umol/L; normal range 5.1-28.0) | 9.95(5.03) |
| Decreased | 3(16.67) |
| Blood urea nitrogen (mmol/L; normal range 2.9-8.2) | 4.18(1.22) |
| Decreased | 1(5.56) |
| Serum creatinine (umol/L; normal range 44.0-123.0) | 70.44(17.55) |
| Decreased | 1(5.56) |
| Creatine kinase-MB isoenzyme activity (ng/ml; normal range 0.0-5.0) | 2.72(0.30) |
| Lactate dehydrogenase (U/L; normal range 109.0-245.0) | 254.99(63.35) |
| Increased | 8(44.44) |
| Myoglobin (ng/mL; normal range 0.0-146.0) | 30.41(1.36) |
| Glucose (mmol/L; normal range 3.9-6.1) | 4.60(0.65) |
| Increased | 1(5.56) |
| Decreased | 1(5.56) |
| **Infection-related biomarkers** |  |
| Procalcitonin (ng/mL; normal range <0.5) | <0.5 |
| C-reactive protein (mg/L; normal range 0-10) | 5.03(0.12) |

\* Changes (i.e., above or below the normal range) were summarized as number of increase/decrease (% of involved individuals.)

## Discussions

To our knowledge, this is the first descriptive study specifically focusing on a group of asymptomatic individuals with SARS-CoV-2 infection. We found that in Tibetan population, asymptomatic carriers were mainly young or middle-aged healthy individuals, with their infections disappeared after antiviral treatment. Importantly, although no clinical symptoms, many of them have typically viral pneumonia-like changes in chest CT images and abnormal laboratory measures.

The possible abnormal findings on chest CT images among asymptomatic carriers got support from one previous report[2]. However, the common abnormalities in blood tests was never described before[2-4]. Also, it is notable that there is a considerable proportion of asymptomatic carriers among our study population (21.7%), which was much higher than the rate reported previously (5%)[5]. This may due to a high screening rate for individuals with exposure history in Ganzi area. Alternatively, the genetic variations between Han Chinese and Tibetans may be attributable for these differences.

The major merits of our study include a sufficient follow-up period and a high coverage rate of all individuals with SARS-CoV-2 infection in study area. Our finding indicates a potentially big number of SARS-CoV-2 asymptomatic carriers in prevalent area, highlighting a necessity of screening individuals with close contact of infected patients, for a better control on the spread of SARS-CoV-2 infection.

## Data Availability

The data used in this study are compiled in the COVID-19 database at Sichuan Univeristy and are available for review on site, on request.

## Acknowledgements

We thank Yao Hu, Junren Wang, Jingwei Jiang, Tingxi Zhu, Yilong Chen, Xiaoxi Zeng, Chunyang Li, Yajing Sun, Yonghong Gu, Chao Zhang, for data extraction and data cleaning. None of these individuals received compensation for their contributions.

